# Dose-Related Effects of Ketamine for Antidepressant-Resistant Symptoms of Posttraumatic Stress Disorder in Veterans and Active Duty Military: A Double-blind, Randomized, Placebo-Controlled Multi-Center Clinical Trial

**DOI:** 10.1101/2021.04.30.21256273

**Authors:** Chadi G. Abdallah, John D. Roache, Ralitza Gueorguieva, Lynnette A. Averill, Stacey Young-McCaughan, Paulo R. Shiroma, Prerana Purohit, Antoinette Brundige, William Murff, Kyung-Heup Ahn, Mohamed A. Sherif, Eric J. Baltutis, Mohini Ranganathan, Deepak D’Souza, Brenda Martini, Steven M. Southwick, Ismene L. Petrakis, Rebecca R. Burson, Kevin B. Guthmiller, Argelio L. López-Roca, Karl A. Lautenschlager, John P. McCallin, Matthew B. Hoch, Alexandar Timchenko, Sergio E. Souza, Charles E. Bryant, Jim Mintz, Brett T. Litz, Douglas E. Williamson, Terence M. Keane, Alan L. Peterson, Consortium to Alleviate PTSD, John H. Krystal

## Abstract

**Background:** This study tested the efficacy of repeated intravenous ketamine doses to reduce antidepressant-resistant symptoms of posttraumatic stress disorder (PTSD).

**Methods:** Veterans and service members with PTSD (n=158) who failed previous antidepressant treatment were randomized to 8 infusions administered twice weekly of intravenous placebo (n=54), low dose (0.2mg/kg; n=53) or standard dose (0.5mg/kg; n=51) ketamine. Participants were assessed at baseline, during treatment, and for 4 weeks after their last infusion. Primary analyses used mixed effects models. The primary outcome measure was the self-report PTSD Checklist for *DSM-5* (PCL-5), and secondary outcome measures were the Clinician-Administered PTSD Scale for *DSM-5* (CAPS-5) and the Montgomery Åsberg Depression Rating Scale (MADRS).

**Results:** There were no significant group-by-time interactions for PTSD symptoms measured by the PCL-5 or CAPS-5. The standard dose ketamine significantly reduced symptoms after the first infusion, while the low dose showed significant symptom reduction after the last infusion and at the 4-week follow-up. The standard ketamine dose also significantly ameliorated depression measured by the MADRS. Ketamine produced dose-related dissociative and psychotomimetic effects, which returned to baseline within 2 hours and were less pronounced with repeated administration. There was no evidence of differential treatment discontinuation by ketamine dose, consistent with good tolerability.

**Conclusions:** This clinical trial failed to find a significant dose-related effect of ketamine on PTSD symptoms. Secondary analyses suggested that the low dose reduced PTSD symptoms and the standard dose exerted rapid antidepressant effects. Further studies are needed to determine the role of ketamine in PTSD treatment.

**ClinicalTrials.gov identifier:** *NCT02655692*

## INTRODUCTION

Posttraumatic stress disorder (PTSD) is a debilitating illness with limited pharmacotherapy options (1-3). Patients are often treated with monoaminergic antidepressants and off-label combinations of other pharmacotherapies, most of which have inadequate evidence for efficacy in PTSD treatment (4,5). Moreover, meta-analytic studies show only small differences between pharmacotherapy and placebo (6), particularly in veterans suffering from PTSD (7).

Ketamine, an antagonist of N-methyl-D-aspartate (NMDA) glutamate receptors, is a rapid-acting antidepressant with a novel mechanism of action (8,9). In a primarily civilian sample, a pioneering proof-of-concept study (n=41) showed rapid reduction in PTSD and depression symptoms 24 hours post a single standard ketamine dose (0.5 mg/kg intravenously over 40 minutes) compared to midazolam (10). The standard dose ketamine administered 3 times per week for 2 weeks also was recently reported to significantly reduce PTSD symptoms compared to midazolam in a randomized controlled pilot study (n=30) (11). Additionally, an uncontrolled, open label small study (n=15) in subjects with comorbid PTSD and major depressive disorder (MDD) reported reductions in both PTSD and MDD symptoms following treatment with 6 standard ketamine intravenous infusions over a 2-week period (12). But another small open-label study (n=10) in individuals with comorbid PTSD and MDD found no significant effects on PTSD symptoms 24 hours following single standard ketamine infusion (13). Moreover, a randomized controlled clinical trial (n=40) found no significant effects of a single standard infusion of ketamine compared to midazolam in 4 groups of individuals with comorbid PTSD and/or chronic pain (14). Together, previous studies suggest the potential utility of ketamine in treating PTSD symptoms. However, the evidence is mixed and, to date, only the standard dose ketamine has been tested. These findings underscore the need for larger, and more definitive, placebo-controlled trials to determine the efficacy of ketamine in treating PTSD symptoms.

This study investigated the efficacy of the standard (0.5 mg/kg) and a low dose (0.2 mg/kg) of ketamine in a double-blind, randomized, placebo-controlled clinical trial in veterans and active duty service members with antidepressant-resistant PTSD symptoms (15). Considering that the effects of ketamine are short-lived following single infusion (10), we tested the efficacy and durability of repeated ketamine. At the time the study was designed, the evidence suggested that the rapid acting antidepressant ketamine administered twice per week is comparable to 3 times per week (16). Hence, we opted to administer the study drug twice weekly for a total of 8 infusions. Moreover, previous data suggested that benzodiazepine treatment may worsen PTSD outcomes (17), therefore we opted for inactive placebo control in this repeated administration study. Considering that the dissociative symptoms of ketamine are dose dependent, we anticipated that the low dose ketamine will enhance the functional blinding. Although our primary hypothesis was focused on PTSD symptoms, we also evaluated the efficacy of ketamine against the depressive symptoms that are highly comorbid in the study population (18). Moreover, this study also evaluated the dissociative and psychotomimetic effects of ketamine as well as other adverse events to determine the safety of repeated ketamine in patients with PTSD, an illness at times characterized by dissociative pathology (19).

We hypothesized that a standard dose of ketamine would exert a rapid reduction in PTSD symptoms, compared to placebo, at 24 hours post-first infusion, and that this therapeutic benefit would persist through the end of treatment. Moreover, we anticipated that the repeated dosing would maintain the therapeutic response during the 4-week follow-up period.

## METHODS

### Study Design

Full details of the study methods were previously reported (15). In summary, this multi-center double-blind randomized controlled trial enrolled veterans and active duty service members with antidepressant-resistant PTSD symptoms. This study was part of the Consortium to Alleviate PTSD (CAP), an initiative supported jointly by the US Department of Defense and the US Department of Veterans Affairs. All study procedures were approved and monitored by an Institutional Review Board at each study site as well as the CAP Data and Safety Monitoring Board and the US Army Medical Research and Development Command Human Research Protection Office. All participants completed an informed consent process prior to enrollment.

Between September 2016 and March 2020, participants were randomized to three parallel study arms: 1) Placebo (normal saline); 2) Low dose (ketamine 0.2 mg/kg); 3) Standard dose (ketamine 0.5 mg/kg). Eight 40-minute intravenous infusions of the study drug were administered twice weekly. Participants were assessed weekly for 4 weeks following the last infusion. The Clinician-Administered PTSD Scale for *DSM-5* (CAPS-5) was used to confirm the PTSD diagnosis and to assess severity of symptoms at baseline, at the end of treatment, and the end of follow-up. The self-reported PTSD Checklist for *DSM-5* (PCL-5) and clinician-administered Montgomery-Åsberg Depression Rating Scale (MADRS) assessed PTSD and depressive symptoms, respectively. These ratings were administered prior to each infusion, at 24 hours post-first and post-last infusions, and weekly during follow-up. The dissociative and psychotomimetic effects of ketamine were assessed using the Clinician-Administered Dissociative State Scale (CADSS) and the Positive and Negative Syndrome Scale (PANSS), respectively. These were administered at 30 minutes and 120 minutes from the start of the study drug infusion. At the end of treatment period, participants were asked to guess which study drug they were on and how confident they are in their guess.

Participants who did not respond were offered a single administration of open label, standard dose ketamine; their follow-up data were not included in the durability of effect analyses. Response was defined as at least 25% improvement in CAPS-5 scores following treatment as compared to baseline (15). The study target was to randomize 198 subjects. However, due to the restrictions in direct patient care imposed by the COVID-19 pandemic, the study was prematurely closed to new enrollment.

### Study Criteria

The study enrolled veterans and service members between the age of 18 and 70 years (15). Participants met the following criteria: 1) were diagnosed with PTSD based on the structured CAPS-5 interview; 2) had CAPS-5 score of 23 or higher (i.e., moderate to severe); 3) had a history of nonresponse to at least 1 adequate trial of FDA approved antidepressant, as determined by the Massachusetts General Hospital Antidepressant Treatment History Questionnaire (MGH-ATRQ); 4) were unmedicated or were stable on an antidepressant for at least 4 weeks or PTSD-focused psychotherapy for at least 6 weeks; 5) if female, were not pregnant or breastfeeding and were on a medically acceptable contraceptive method; 6) were able to read, write, and provide written informed consent in English; 7) did not have psychotic disorder or features, or manic or mixed episodes; 8) did not have an unstable medical condition; 9) had no suicidal or homicidal risk meriting crisis intervention; 10) had no severe brain injury; 11) had no moderate or severe substance use disorder within 3 months – except for mild-moderate alcohol use disorder with negative breathalyzer; 12) were not currently using monoamine oxidase inhibitors, memantine, or long-acting benzodiazepines and had no known sensitivity to ketamine; and 13) had resting blood pressure higher than 90/60 and lower than 150/90 mmHg, and heart rate higher than 45/min and lower than 100/min.

### Statistics

Power calculation and analysis plans were previously reported (15) and are further detailed in the online Supplemental Information. Briefly, the PCL-5 was considered the primary outcome, while CAPS-5 and MADRS were considered secondary measures. The primary analysis used mixed effects models, with group, time, and group-by-time effects. The secondary analyses examined the rapid and sustained effects of ketamine compared to placebo, at 24 hours post-first and post-last infusion, respectively, by focused contrasts in the mixed models. Sustainability of the effects of ketamine on PTSD symptoms was assessed with similar mixed models for PCL-5 and CAPS-5 during the follow-up period; considering that this analysis included only the responders (non-responders received open label ketamine), we covaried for pretreatment symptom severity. However, there was no difference in pretreatment severity and the results are similar without covarying for severity. The dissociative and psychotomimetic effects were examined using comparable mixed models for CADSS and PANSS, while adding interval (30 minutes vs. 120 minutes) and appropriate interactions to the models.

## RESULTS

A total of 262 individuals were consented and assessed for eligibility. Of these individuals, 158 were found eligible, randomized, began treatment, and included in the study analysis. The CONSORT flow diagram is provided in the Supplement. The demographics of the randomized participants are provided in Table 1. Participants had moderate to severe symptoms of PTSD and depression (see PCL-5 and MADRS in Table 1). Among the patients randomized for treatment, the discontinuation rate was similar across the 3 groups (placebo: 19%, low: 15%, standard: 16%; χ^2^(2) = 0.3, p = 0.88). The majority of participants believed they were on low dose ketamine (standard: 19%; low: 57%; placebo: 24%). The percent of participants who correctly guessed the study drug differed between groups (standard: 37%; low: 78%; placebo: 64%; χ^2^(4) = 62.7, p < 0.0001). Only 29% of participants were confident of their guess, with no significant difference in the confidence between those who correctly and incorrectly guessed the study drug (χ^2^(1) = 1.0, p = 0.31).

**Table 1.** Demographics and Clinical Characteristics.

|  | Standard Dose | Low Dose | Placebo |  |
| --- | --- | --- | --- | --- |
|  | n=51<br>0.5 mg/kg | n=53<br>0.2 mg/kg | n=54<br>placebo | p value |
| Sex – male (%) | 38 (74.5) | 43 (81.1) | 40 (74.1) | 0.63 |
| Mean age (SD) | 43.2 (12.7) | 45.2 (11.2) | 42.0 (10.8) | 0.37 |
| White - Non-Hispanic | 29 (56.9) | 28 (52.8) | 37 (68.5) | 0.57 |
| Black | 7 (13.7) | 7 (13.2) | 6 (11.1) |  |
| Hispanic | 12 (23.5) | 11 (20.8) | 8 (14.8) |  |
| Other | 3 (5.9) | 7 (13.2) | 3 (5.6) |  |
| < 12th grade, high school | 6 (11.8) | 5 (9.4) | 7 (13.0) | 0.66 |
| Some college | 20 (39.2) | 17 (32.1) | 15 (27.8) |  |
| College degree | 21 (41.2) | 25 (47.2) | 22 (40.7) |  |
| Some graduate | 4 (7.8) | 6 (11.3) | 10 (18.5) |  |
| Duty Status - Veteran | 34 (66.7) | 36 (67.9) | 37 (68.5) | 0.98 |
| Active duty | 17 (33.3) | 17 (32.1) | 17 (31.5) |  |
| Army | 29 (56.9) | 32 (60.4) | 33 (61.1) | 0.59 |
| Navy | 8 (15.7) | 10 (18.9) | 10 (18.5) |  |
| Air Force | 4 (7.8) | 6 (11.3) | 7 (13.0) |  |
| Marine | 10 (19.6) | 5 (9.4) | 4 (7.4) |  |
| # Deployments |  |  |  | 0.12 |
| 0 | 19 (37.3) | 20 (37.7) | 17 (31.5) |  |
| 1 | 15 (29.4) | 7 (13.2) | 11 (20.4) |  |
| 2 | 14 (27.5) | 13 (24.5) | 13 (24.1) |  |
| 3+ | 3 (5.9) | 13 (24.5) | 13 (24.1) |  |
| Years military service | 11.2 (7.5) | 12.0 (8.6) | 10.9 (8.3) | 0.77 |
| PCL-5 | 47.5 (14.5) | 46.6 (17.7) | 48.6(12.8) | 0.80 |
| CAPS-5 | 25.1 (13.6) | 25.8 (17.7) | 29.7 (14.6) | 0.38 |
| MADRS | 27.8 (9.3) | 27.8 (10.3) | 28.2 (8.4) | 0.97 |

**Abbreviations:** PCL-5: PTSD Checklist for *DSM-5*; CAPS-5: Clinician-Administered PTSD Scale for *DSM-5*; MADRS: Montgomery-Åsberg Depression Rating Scale.

### Effects of Ketamine on Primary Outcome

The primary analysis found no significant treatment-by-time interactive effect on the self-reported PCL-5 scores (F_(18,137)_ = 1.1, p = 0.38; Fig. 1A). There was a time effect (F_(9,133)_ = 37.1, p < 0.0001) wherein PCL-5 scores improved across all treatment groups, with no treatment main effect (F_(2,148)_ = 1.8, p = 0.17). The effect sizes are reported in Table 2. Secondary analyses showed rapid reduction in PCL-5 score 24 hours after the initial dose in the standard dose group, compared to placebo (mean difference (±SEM) = 6.6 (±3.1), t(149)=2.1, p = 0.04). However, compared to placebo, these changes were not significant at the end of treatment (mean difference (±SEM) = 5.0 (±3.4), t(147)=1.5, p = 0.14). In contrast, there was a significant effect of the low dose compared to placebo at the end of treatment (mean difference (±SEM) = 6.4 (±3.3), t(147)=2.0, p = 0.05), but not after the first infusion (mean difference (*±SEM*) = 3.3 (*±3*.*1)*, t(149)=0.3, p = 0.29). There were no differences between standard and low doses of ketamine (p > 0.1).

**Figure 1.**
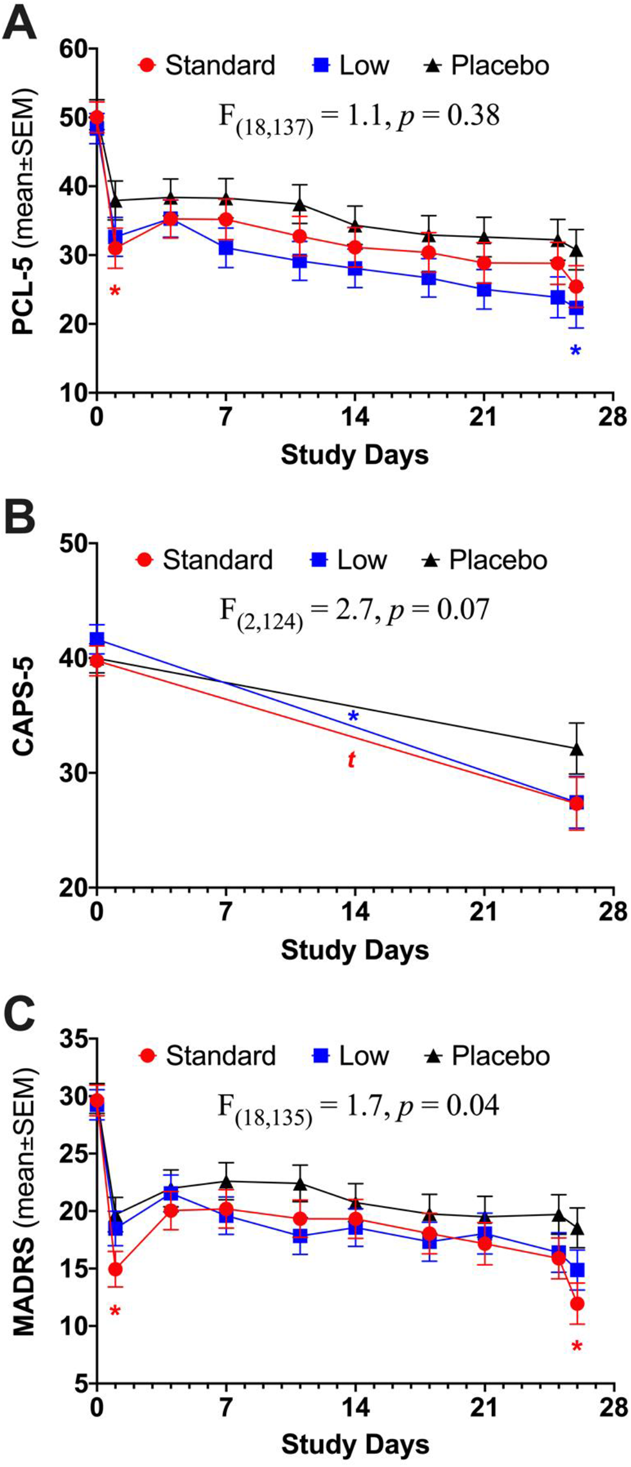
The effects of ketamine on posttraumatic stress disorder (PTSD) and depression symptoms. **A**. The PTSD Checklist for *DSM-5* (PCL-5) scores were significantly reduced over the treatment period but did not differ between the treatment groups. Secondary analysis showed rapid reduction in PCL-5 following standard dose ketamine compared to placebo at 24 hours post first infusion (red * on Day 1). There was also significant reduction in PCL-5 at 24 hours post last infusion in low dose ketamine compared to placebo (blue * on Day 26). **B**. There was a trend effects of ketamine on the Clinician-Administered PTSD Scale for DSM-5 (CAPS-5), reflecting significant reduction in CAPS-5 in the low dose (blue *) and a trend in the standard dose (red t) over the treatment period compared to placebo. **C**. The Montgomery-Åsberg Depression Rating Scale (MADRS) scores were significantly reduced over the treatment period. This MADRS reduction differed between the treatment groups. There was significant improvement in depression symptoms at 24 hours and end of treatment in the standard dose ketamine compared to placebo (red * on Day 1 and 26). There was no significant improvement in depression symptoms following low dose ketamine compared to placebo. *<u>Notes</u>*: Assessments collected prior to each study drug infusion, except on Day 1 and Day 25, which were collected 24 hours post-first and post-last infusions, respectively.

**Table 2.** Treatment Effect Sizes (Cohen d’)

|  | Rapid<br>24h post first infusion | End of Treatment<br>24h post last infusion | Follow-up<br>4 weeks post last infusion |
| --- | --- | --- | --- |
| PCL-5 |  |  |  |
| Placebo | 0.75 | 1.13 | 1.01 |
| Ketamine 0.2mg/kg | 0.93 | 1.53 | 1.31 |
| Ketamine 0.5mg/kg | 0.96 | 1.61 | 0.89 |
| CAPS-5 |  |  |  |
| Placebo |  | 0.66 | 1.01 |
| Ketamine 0.2mg/kg |  | 1.01 | 1.68 |
| Ketamine 0.5mg/kg |  | 1.15 | 1.41 |
| MADRS |  |  |  |
| Placebo | 1.25 | 1.14 | 0.89 |
| Ketamine 0.2mg/kg | 1.06 | 1.35 | 1.06 |
| Ketamine 0.5mg/kg | 1.53 | 1.81 | 0.82 |

PCL-5 = PTSD Checklist for *DSM-5*; CAPS-5 = Clinician-Administered PTSD Scale for *DSM-5*; MADRS = Montgomery Åsberg Depression Rating Scale.

The percentage of responders (i.e., ≥ 25% improvement in PCL-5; Fig. 2) at 24h post-first infusion is higher in the two active groups (47% on ketamine standard dose, 47% on ketamine low dose) than in the placebo group (33% on placebo), but the difference is not significant in the logistic model (Fig. 2). The odds of reaching responder status on active treatment are more than 80% higher than the odds on placebo, but the effects are not statistically significant [OR=1.88, 95% CI: (0.84,4.22) for standard dose vs. placebo, OR=1.82, 95% CI: (0.82,4.05) for low dose vs. placebo]. Similarly, the percentage of responders at 24h post last infusion is higher in the two active groups (63% on ketamine standard dose, 62% on ketamine low dose) than in the placebo group (52% on placebo), but the difference is not significant in the logistic model. The odds of reaching responder status on active treatment are more than 50% higher than the odds on placebo, but the effects are not statistically significant [OR=1.61, 95% CI: (0.73,3.53) for standard dose vs. placebo, OR=1.55, 95% CI: (0.71,3.37) for low dose vs. placebo]. Examining the 4 clusters of PTSD separately yielded comparable results to the total PCL-5 (i.e., no treatment-by-time interactions with all p values > 0.05). Additional exploratory analyses can be found in the Supplements.

**Figure 2.**
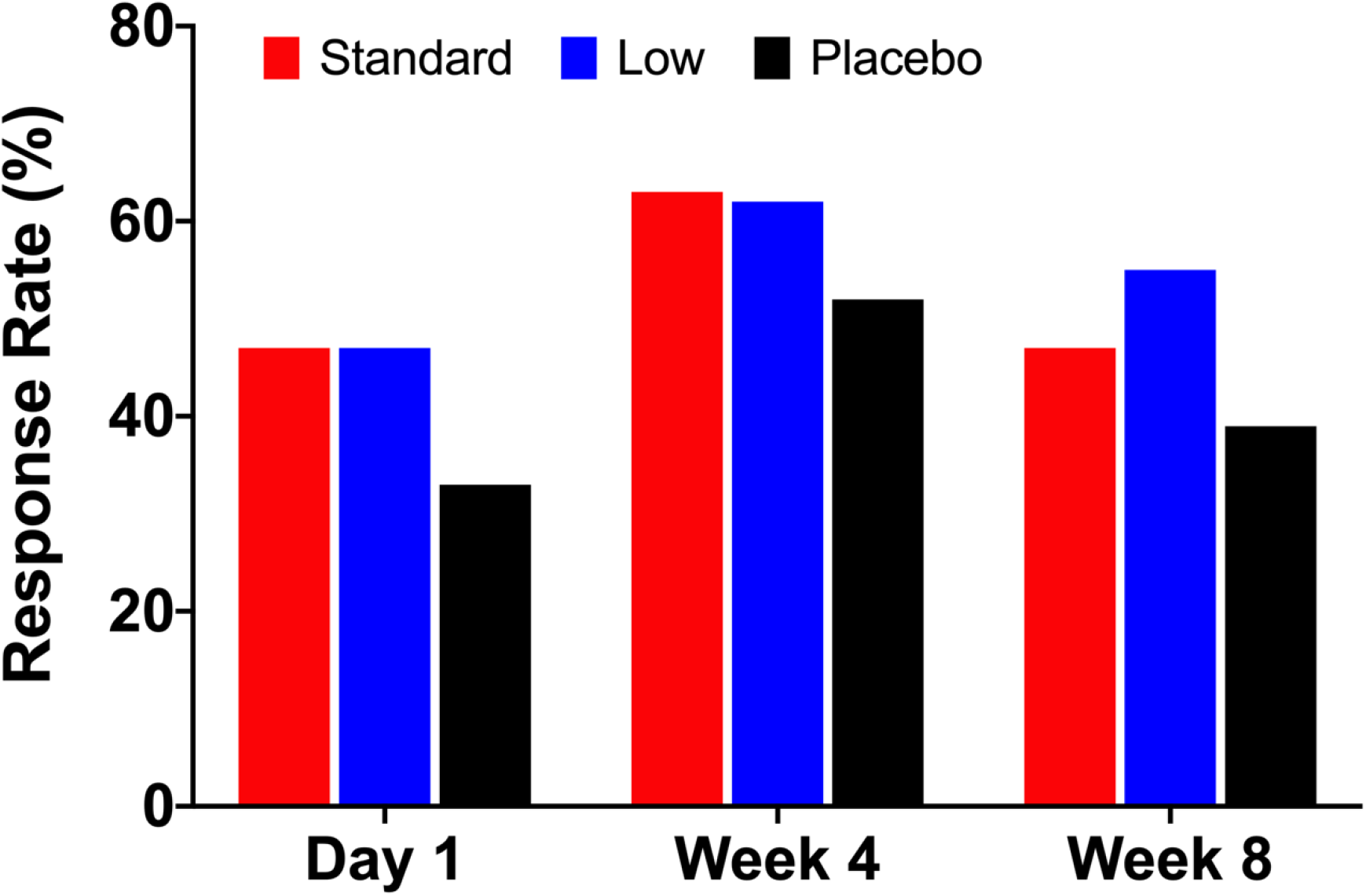
The response rate during and following treatment. There was no significant difference in response rate (i.e., 25% or more improvement in the PTSD Checklist for *DSM-5* (PCL-5) scores) at 24h post first infusion (Day 1), 24h post last infusion (Week 4), and 4 weeks post last infusion (Week 8).

### Effects of Ketamine on Secondary Outcomes

The primary analysis showed a nonsignificant trend of treatment-by-time interactive effect on the clinician-administered CAPS-5 scores (F_(2,124)_ = 2.7, p = 0.07; Fig. 1B). There was a time effect (F_(1,124)_ = 103.4, p < 0.0001) but no treatment main effect (F_(2,145)_ = 0.8, p = 0.46). Compared to placebo, the secondary analyses showed significant reduction in CAPS-5 at the end of treatment in the ketamine low dose (mean difference (±SEM) = 6.0 (±2.7), t(124)=2.2, p = 0.03), but not the ketamine standard dose group (mean difference (±SEM) = 4.7 (±2.8), t(124)=1.7, p = 0.09). There were no differences between standard and low doses of ketamine (p > 0.1).

In contrast, ketamine had significant dose-related effects on depression symptoms. In the analysis of MADRS data, the mixed model showed a significant treatment-by-time interaction (F_(18,135)_ = 1.7, p = 0.04, Fig 1C) and a time effect (F_(9,133)_ = 35.0, p < 0.0001) but no treatment main effect (F_(2,150)_ = 1.3, p = 0.28). Secondary analyses showed rapid reduction in MADRS at 24 hours post-first infusion in the standard ketamine dose group: standard vs. placebo (mean difference (±SEM) = 4.6 (±1.9), t(148)=2.5, p = 0.02) and standard vs. low dose (mean difference (±SEM) = 3.9 (±1.9), t(149)=2.1, p = 0.04). At the end of treatment, there was significant MADRS reduction in the ketamine standard dose compared to placebo (mean difference (±SEM) = 6.4 (±2.2), t(140)=2.9, p = 0.004) but not compared to the low dose group (mean difference (±SEM) = 3.3 (±2.2), t(141)=1.5, p = 0.14). There were no differences between ketamine low dose and placebo (p > 0.1).

### Durability of the Ketamine Effects

After the end of the 4-week treatment period, participants in the standard dose (n=13; 25%), low dose (n=18; 34%), and placebo groups (n=25; 46%) with less than 25% improvement on the CAPS-5 were considered nonresponders and were offered open-label, standard dose ketamine (Chi-sq(2)=5.0, p=0.08). Excluding those with open-label, the durability mixed model analysis showed a nonsignificant trend for a treatment-by-time interactive effect on the PCL-5 scores (F_(6,68)_ = 2.0, p = 0.08). There was a time effect (F_(3,68)_ = 6.5, p < 0.001) but no treatment main effect (F_(2,68)_ = 2.3, p = 0.11). At 4 weeks posttreatment, PCL-5 scores remained significantly lower in the ketamine low dose compared to placebo (mean difference (±SEM) = 15.3 (±5.8), t(68)=2.6, p = 0.01) and standard dose groups (mean difference (±SEM) = 9.8 (±4.8), t(68)=2.0, p = 0.05). There were no differences between placebo and ketamine standard dose (p > 0.1).

Examining the durability of ketamine effects on CAPS-5 scores, the mixed model showed main time effect (F_(2,71)_ = 54.7, p < 0.0001) but no treatment (F_(2,127)_ = 1.4, p = 0.26) or treatment*group effects (F_(4,70)_ = 1.9, p = 0.13). At 4 weeks posttreatment, CAPS-5 scores were significantly reduced in the ketamine low dose compared to the placebo (mean difference (±SEM) = 8.4 (±3.7), t(65)=2.2, p = 0.03) but not the standard dose group (mean difference (±SEM) = 2.7 (±3.4), t(63.7)=0.8, p = 0.43). The CAPS-5 reduction in the ketamine standard dose was not significant compared to placebo (mean difference (±SEM) = 5.7 (±3.7), t(64.9)=1.6, p = 0.13). Similarly, there was a main effect of time on MADRS (F_(3,70)_ = 5.8, p = 0.001), but no treatment (F_(2,70)_ = 1.3, p = 0.29) or treatment*group effects (F_(6,70)_ = 0.8, p = 0.55). At 4 weeks posttreatment, MADRS scores were significantly lower in the ketamine low dose compared to placebo (mean difference (±SEM) = 7.9 (±3.5), t(70)=2.3, p = 0.03), but no differences between other groups (p > 0.05).

### Adverse Effects of Ketamine

Examining dissociative effects of ketamine as measured by CADSS, the mixed model analysis showed significant effect of treatment (F_(2,147)_ = 20.2, p < 0.0001), with dose-dependent ketamine-induced dissociative symptoms (Fig. 3A). As expected, there was a significant treatment by interval (i.e., during vs. post) interaction (F_(2,803)_ = 102.1, p < 0.0001), with the ketamine-induced dissociative symptoms observed during treatment dissipating 80 minutes after the 40-minute infusion was complete (Fig. 3A). We also found time effects on CADSS (F_(7,133)_ = 8.6, p < 0.0001), with a reduction of dissociative symptoms observed over the 8-infusion treatment period (Fig. 3B).

**Figure 3.**
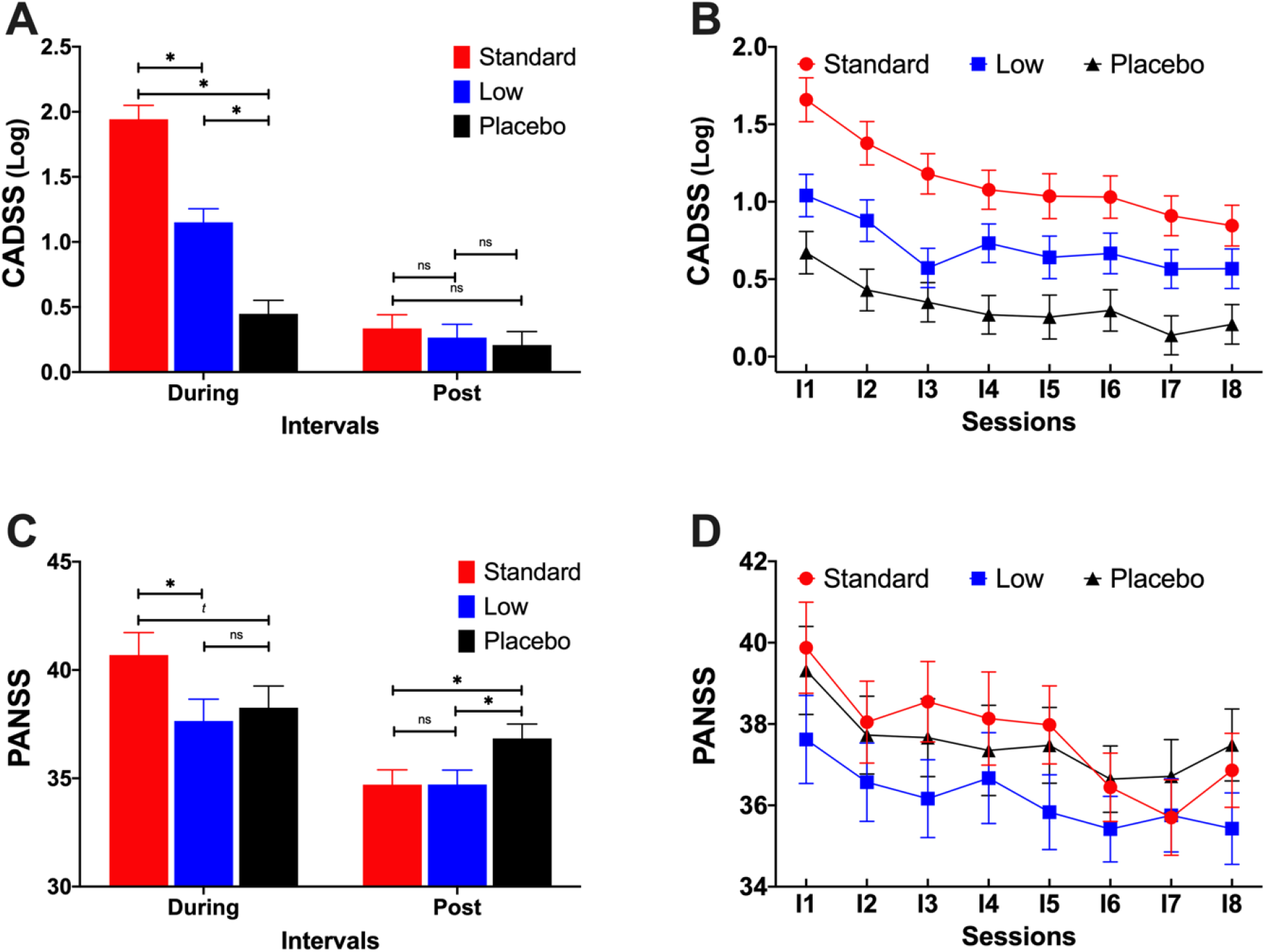
The dissociative and psychotomimetic effects of ketamine treatment in patients with posttraumatic stress disorder (PTSD). **A**. There was a dose-dependent, ketamine-induced increase in the Clinician-Administered Dissociative State Scale (CADSS) scores at 30 minutes from the start of the infusion (During). This ketamine-induced dissociation symptoms returned to placebo levels at the 120 minutes time point (Post). **B**. There was a significant time effect on CADSS scores, indicating reduction in the dissociative symptoms with repeated treatment, from the first (I1) to last infusion (I8). **C**. The standard dose induced increase in the Positive and Negative Syndrome Scale (PANSS) scores at 30 minutes from the start of the infusion (During). This ketamine-induced psychotomimetic effect improved at the 120 minutes time point (Post). **D**. There was a significant time effect on PANSS scores, indicating reduction in the psychotomimetic symptoms with repeated treatment, from the first (I1) to last infusion (I8). *<u>Abbreviations</u>*: **ns** indicates p values > 0.10; ***t*** indicates p values < 0.10 * indicates p values < 0.05; Standard = ketamine 0.5 mg/kg; Low = ketamine 0.2 mg/kg.

Examination of the psychotomimetic effects using the PANSS showed results comparable to the CADSS findings, with significant treatment*interval interactive effects (F_(2,148)_ = 13.9, p < 0.0001; Fig. 3C). There was also significant time effect (F_(7,128)_ = 5.1, p < 0.0001; Fig. 3D), but no effect of treatment (F_(2,148)_ = 1.2, p = 0.3). Follow-up analyses showed ketamine-induced psychotomimetic symptoms observed during infusion significantly improved by 120 minutes.

The majority of participants (n=137, 87%) reported at least one adverse event (AE), with a total of 402 AEs during the study. Of the 402 AEs, 273 occurred during the treatment infusion period, and 162 of them were considered at least “possibly” related to treatment. There were 13 treatment-related AEs that occurred in more than 2 participants, and those are displayed in the Supplemental Table S1. AEs most likely associated with one of the active ketamine doses were agitation, anxiety, irritability, and constipation, which occurred infrequently in the ketamine groups and not at all in the placebo group. Notably, nightmare occurrence was comparable across groups, while headache was more common in the low dose ketamine group. Nausea or other gastrointestinal disturbance occurred equally frequently in all groups including placebo.

## DISCUSSION

In the largest sample and longest treatment duration studied to date, this clinical trial failed to demonstrate the efficacy of 4 weeks of twice-weekly ketamine infusions to treat antidepressant-resistant PTSD symptoms in veterans and active duty military. This was true despite observing a significant antidepressant effect of ketamine in these patients who had considerable depressive symptoms at baseline. Nonetheless, a set of *a priori* planned secondary analyses support the need for further research to determine the optimal dose and frequency of ketamine in treating PTSD. First, consistent with reported pilot findings (10), the standard ketamine dose (0.5 mg/kg) exerted rapid reduction in PTSD symptoms at 24h post first infusion. Moreover, low dose ketamine appears to produce a gradual reduction in PTSD symptom severity, evident after the second infusion and was significantly different compared to placebo at the end of treatment. Notably, the therapeutic benefit of low dose ketamine relative to placebo was significant at 4 weeks post last treatment as measured by the PCL-5, the CAPS-5 and the MADRS scales.

A major challenge in ketamine research is the potential for functional unblinding due to the distinguishing acute dissociative effects of ketamine (20). The concern is that functional unblinding in ketamine studies may lead to low placebo effects and exaggerated response to the index study drug. Midazolam, a benzodiazepine, has been previously proposed as putative active control in ketamine studies (21). However, the potential negative effects of benzodiazepine on PTSD are previously documented and should be avoided as control in PTSD treatment studies (17). In the current study, the majority of participants incorrectly guessed they were on low dose ketamine. Participants also had low confidence in their guesses, regardless whether they correctly or incorrectly guessed their study drug. Furthermore, only 37% of participants in the standard dose correctly guessed their treatment. Together, these data suggest that the use of low dose ketamine may have enhanced the blinding of the standard dose, and presumably increased participants expectation in the placebo dose. In fact, we observed increased instead of reduced placebo effects. The effect sizes of both ketamine doses on PCL-5 scores were large and in the predicted range (0.93-1.61), but the effect size of placebo was larger than expected (0.75-1.13), resulting in only small effects of ketamine treatment relative to placebo. Unfortunately, failed clinical trials due to high placebo response are not uncommon in this field (22). Various factors may have contributed to the high placebo response, including the repeated invasive medical interventions of intravenous infusion twice per week for 4 weeks requiring 2-3 weekly visits approximately 4 hours each including comprehensive assessments and appropriate supportive milieu of attending to the participant needs during the study period (e.g., booking transportations, meals, etc.).

Depression symptoms, which were substantial in the current cohort, are commonly associated with PTSD, and have been reported to respond relatively poorly to traditional antidepressants. For example, in the VAST-D study, depressed patients with PTSD had poorer overall outcomes than depressed patients without PTSD (23). In the current study, ketamine showed significant effect in the mixed model and in the secondary analyses, reflecting rapid antidepressant effects on Day 1 and at the end of treatment in the standard dose group. Consistent with the depression literature (24,25), the low dose ketamine had no rapid antidepressant effect during treatment. However, at the end of the 4-week follow-up, participants in the low dose group were found to have reduced depressive symptoms compared to placebo, suggesting delayed relapse of depression in the low dose group.

The current study does not support the pilot findings by Feder and colleagues (11), which reported significant effects of standard dose ketamine on PTSD symptoms. Several differences between the trials may have contributed to the differing results. Our study was in military population, while the previous report was primarily in civilians. Furthermore, our participants were mostly males with only 23% females compared to the previous study with 77% females. The sex differences may have played a role in the differing outcomes. Considering the low number of females per group, we were not able to statistically assess the effect of sex in our cohort. Other differences in the previous study (11) compared to the current trial, include: 1) smaller cohort of 15 subjects per group, 2) using benzodiazepine as control, 3) no low dose arm, 4) administering the study drugs 3 times per week, 5) treatment was for 2 weeks, and 6) the previous study did not target treatment-resistant PTSD. Notably, the previous study found no rapid effects of ketamine on PTSD or depression symptoms at 24h post first infusion (11). This could be due to the small sample size but it also underscores the challenges of demonstrating the therapeutic effects of standard dose ketamine in PTSD patients (13,14).

Finally, the current study demonstrated the feasibility and short-term safety of repeated intravenous ketamine in a large cohort of patients with PTSD. A major concern in the field was whether adverse effects of repeated ketamine doses would exacerbate PTSD symptoms (15). However, consistent with a recent report (11), the study treatment regimen was found to be well tolerated, showing significant reductions in the dissociative and psychotomimetic symptoms over the treatment period. Moreover, the ketamine-induced dissociative and psychotomimetic effects were transient, returning to normal levels within 2h of starting the ketamine infusion. These dissociative effects were not so severe as to have an impact on retention in treatment over 8 intravenous infusions. Importantly, as has been reported in depressed patients (25), the ketamine-induced dissociative effects were significantly lower during ketamine 0.2 mg/kg compared to the standard dose (0.5 mg/kg) commonly used to treat depression. The latter finding suggests that using lower doses, if they were found efficacious in PTSD, might offer superior tolerability.

In summary, the current study failed to find a significant dose-related effect of ketamine on PTSD symptoms in veterans and military personnel with treatment-resistant symptoms of PTSD. The study found evidence of rapid antidepressant effects in this population. It also highlighted the need to further investigate lower doses of ketamine in the treatment of PTSD. The study provided data supporting the safety and tolerability of repeated ketamine doses in this population. Together, these findings suggest that ketamine may play a role in managing the complex array of symptoms associated with PTSD, particularly in patients who have not responded to prior pharmacotherapies.

## Data Availability

Data are available upon request.

## CONFLICT OF INTEREST

Dr. Abdallah has served as a consultant, speaker and/or on advisory boards for Genentech, Janssen, Psilocybin Labs, Lundbeck, Guidepoint, and FSV7, and as editor of *Chronic Stress* for Sage Publications, Inc.. He also filed a patent for using mTORC1 inhibitors to augment the effects of antidepressants (Aug 20, 2018). Dr. Krystal is a consultant for Aptinyx, Inc., Atai Life Sciences, AstraZeneca Pharmaceuticals, Biogen, Idec, MA, Biomedisyn Corporation, Bionomics, Limited (Australia), Boehringer Ingelheim International, Cadent Therapeutics, Inc., Clexio Bioscience, Ltd., COMPASS Pathways, Limited, United Kingdom, Concert Pharmaceuticals, Inc., Epiodyne, Inc., EpiVario, Inc., Greenwich Biosciences, Inc., Heptares Therapeutics, Limited (UK), Janssen Research & Development, Jazz Pharmaceuticals, Inc., Otsuka America Pharmaceutical, Inc., Perception Neuroscience Holdings, Inc., Spring Care, Inc., Sunovion Pharmaceuticals, Inc., Takeda Industries, Taisho Pharmaceutical Co., Ltd. Dr. Krystal also reports the following disclosures: **<u>Scientific Advisory Board</u>**: Biohaven Pharmaceuticals, BioXcel Therapeutics, Inc. (Clinical Advisory Board), Cadent Therapeutics, Inc. (Clinical Advisory Board), Cerevel Therapeutics, LLC, EpiVario, Inc., Eisai, Inc., Lohocla Research Corporation, Novartis Pharmaceuticals Corporation, PsychoGenics, Inc., RBNC Therapeutics, Inc., Tempero Bio, Inc., Terran Biosciences, Inc. **<u>Stock</u>**: Biohaven Pharmaceuticals, Sage Pharmaceuticals, Spring Care, Inc. **<u>Stock Options</u>**: Biohaven Pharmaceuticals Medical Sciences, EpiVario, Inc., RBNC Therapeutics, Inc., Terran Biosciences, Inc. Tempero Bio, Inc. **<u>Income Greater than $10,000: Editorial Board</u>**: Editor - Biological Psychiatry. **<u>Patents and Inventions</u>**: (1) Seibyl JP, Krystal JH, Charney DS. Dopamine and noradrenergic reuptake inhibitors in treatment of schizophrenia. US Patent #:5,447,948.September 5, 1995. **(2)** Vladimir, Coric, Krystal, John H, Sanacora, Gerard – Glutamate Modulating Agents in the Treatment of Mental Disorders. US Patent No. 8,778,979 B2 Patent Issue Date: July 15, 2014. US Patent Application No. 15/695,164: Filing Date: 09/05/2017. **(3)** Charney D, Krystal JH, Manji H, Matthew S, Zarate C., - Intranasal Administration of Ketamine to Treat Depression United States Patent Number: 9592207, Issue date: 3/14/2017. Licensed to Janssen Research & Development. **(4)** Zarate, C, Charney, DS, Manji, HK, Mathew, Sanjay J, Krystal, JH, Yale University “Methods for Treating Suicidal Ideation”, Patent Application No. 15/379,013 filed on December 14, 2016 by Yale University Office of Cooperative Research. **(5)** Arias A, Petrakis I, Krystal JH. – Composition and methods to treat addiction. Provisional Use Patent Application no.61/973/961. April 2, 2014. Filed by Yale University Office of Cooperative Research. **(6)** Chekroud, A., Gueorguieva, R., & Krystal, JH. “Treatment Selection for Major Depressive Disorder” [filing date 3^rd^ June 2016, USPTO docket number Y0087.70116US00]. Provisional patent submission by Yale University. **(7)** Gihyun, Yoon, Petrakis I, Krystal JH – Compounds, Compositions and Methods for Treating or Preventing Depression and Other Diseases. U. S. Provisional Patent Application No. 62/444,552, filed on January10, 2017 by Yale University Office of Cooperative Research OCR 7088 US01. **(8)** Abdallah, C, Krystal, JH, Duman, R, Sanacora, G. Combination Therapy for Treating or Preventing Depression or Other Mood Diseases. U.S. Provisional Patent Application No. 62/719,935 filed on August 20, 2018 by Yale University Office of Cooperative Research OCR 7451 US01. **<u>On Non-Federal Research Support</u>**: AstraZeneca Pharmaceuticals provides the drug, Saracatinib, for research related to NIAAA grant “Center for Translational Neuroscience of Alcoholism [CTNA-4] Novartis provides the drug, Mavoglurant, for research related to NIAAA grant “Center for Translational Neuroscience of Alcoholism [CTNA-4]. Dr. Gueorguieva discloses royalties from book “Statistical Methods in Psychiatry and Related Fields” published by CRC Press, honorarium as a member of the Working Group for PTSD Adaptive Platform Trial of Cohen Veterans Bioscience and a United States patent application 20200143922 by Yale University: Chekroud, A., Krystal, J., Gueorguieva, R. and Chandra, A. “Methods and Apparatus for Predicting Depression Treatment Outcomes”. Dr. Sherif is a consultant for In Silico Biosciences, Inc. All other co-authors declare no conflict of interest.

## Disclaimer

The views expressed herein are solely those of the authors and do not reflect an endorsement by or the official policy or position of Brooke Army Medical Center, the US Army Medical Department, the US Army Office of the Surgeon General, the Department of the Army, the Department of the Air Force, the Department of Defense, the Department of Veterans Affairs, the National Institutes of Health, or the US Government.

## Additional contributions

The authors would like to thank the individuals who participated in this study for their invaluable contribution. The West Haven site would also like to thank the Emerge Research Program staff and the nurses and lab staff of the Biostudies Unit [Elizabeth O’Donnell RN (Biostudies since October 1989); Angelina Genovese RNC, BSN, MBA (Biostudies since March 1992); Margaret Dion-Marovitz MS, RN (Biostudies since December 2012); Karen E. Prema RN, BSN (Biostudies November 2017 to November 2018)] for the invaluable expertise in conducting this trial and support for its success.

## SUPPLEMENTAL INFORMATION

### Statistical Analyses

Mixed models were constructed to evaluate the effects of posttraumatic stress disorder (PTSD) treatment on the PTSD Checklist for *DSM-5* (PCL-5), Clinician-Administered PTSD Scale for *DSM-5*, Montgomery-Åsberg Depression Rating Scale (MADRS), Clinician-Administered Dissociative State Scale (CADSS), and Positive and Negative Syndrome Scale (PANSS) based on all available observations within individual patients. Treatment group (Standard, Low, Placebo), time (pre each infusion, 24 hours post-first and post-last infusions for PCL-5 and MADRS, baseline and post-last infusion for CAPS-5, during each session for CADSS and PANSS), site and all possible interactions were fit as fixed effects. Alcohol use disorder (AUD) diagnosis was included as a covariate. The CADSS and PANSS models also included interval (30 and 120 minutes) and the interaction between interval and treatment. Subject was the clustering factor. The best-fitting variance-covariance structure in each model was selected based on Bayesian information criterion. Residuals were assessed, and CADSS was log-transformed to correct for positive skewness of this outcome. In the final models, nonsignificant interactions not involving treatment were dropped for parsimony. Focused least square mean comparisons of change from baseline to 24 hours post-first infusion and from baseline to 24 hours post-last infusion within treatment group, and for pairwise differences in change from baseline between treatment groups, were evaluated regardless of the significance of the interactions and main effects. Follow-up data on the PCL-5 and CAPS-5 also were analyzed using mixed models covarying for baseline severity. Individuals with open label treatment were excluded from these analyses. Responder status was calculated as at least 25% improvement from pretreatment on the PCL-5. Missing data were treated as failure (i.e., nonresponder). The primary analyses for the binary outcomes were logistic regressions with treatment, site, and AUD as predictors, and missing treated as failure. The treatment by site interactions were nonsignificant and were dropped for parsimony in the final models.

For the PCL-5 self-assessments, 1.6% of assessments specified incorrect index trauma event referenced for symptom scoring, and 23.6% of assessments were completed without specifying the index trauma. A sensitivity analysis determined that mixed methods ANOVA results were essentially unchanged with or without their inclusion.

### Additional Results

#### Sensitivity Analyses

The main study analyses were repeated to explore the sensitivity of the findings using several post-hoc constructed models. However, none of these post-hoc exploratory analyses affected the main study results (data not shown). These sensitivity analyses included the following:

1. Adjusting for baseline severity based on continuous PCL-5 scores. In this analysis, PCL-5 scores prior to the first infusion were included as covariates in the model.
2. Adjusting for baseline severity based on discrete classification from CAPS-5 scores. In this analysis, participants were classified as having moderate (CAPS-5 ≤ 34) or severe (CAPS-5 > 34) PTSD prior to the first infusion. This severity variable was included as covariate in the model. Interaction between severity and treatment also was tested but showed no significant effects.
3. Accounting for whether or not the index trauma was specified on the self-reported PCL-5. A sensitivity analysis determined that the PCL-5 results did not change when the analysis excluded all PCL-5 self-reports that did not specify the trauma or when the trauma was not consistent across sessions.
4. Restricting analysis to data within 30 days of Session 1. This sensitivity analysis was performed with only major time points for PCL-5, after dropping sessions for which treatment was not competed within 30 days of Session 1.
5. Including Session 1 values as predictor, with data from Session 2 to end-of-treatment as outcome. Consistent with the main study findings, this analysis showed no treatment effects.
6. Using remission instead of response. Remission was defined as CAPS-5 ≤ 10. The results were comparable to the response analyses, with no significant treatment effects.
7. Combining the ketamine groups. Including the standard and low dose as one factor compared to placebo did not affect the main findings. That is, there were no treatment-by-time interactions.

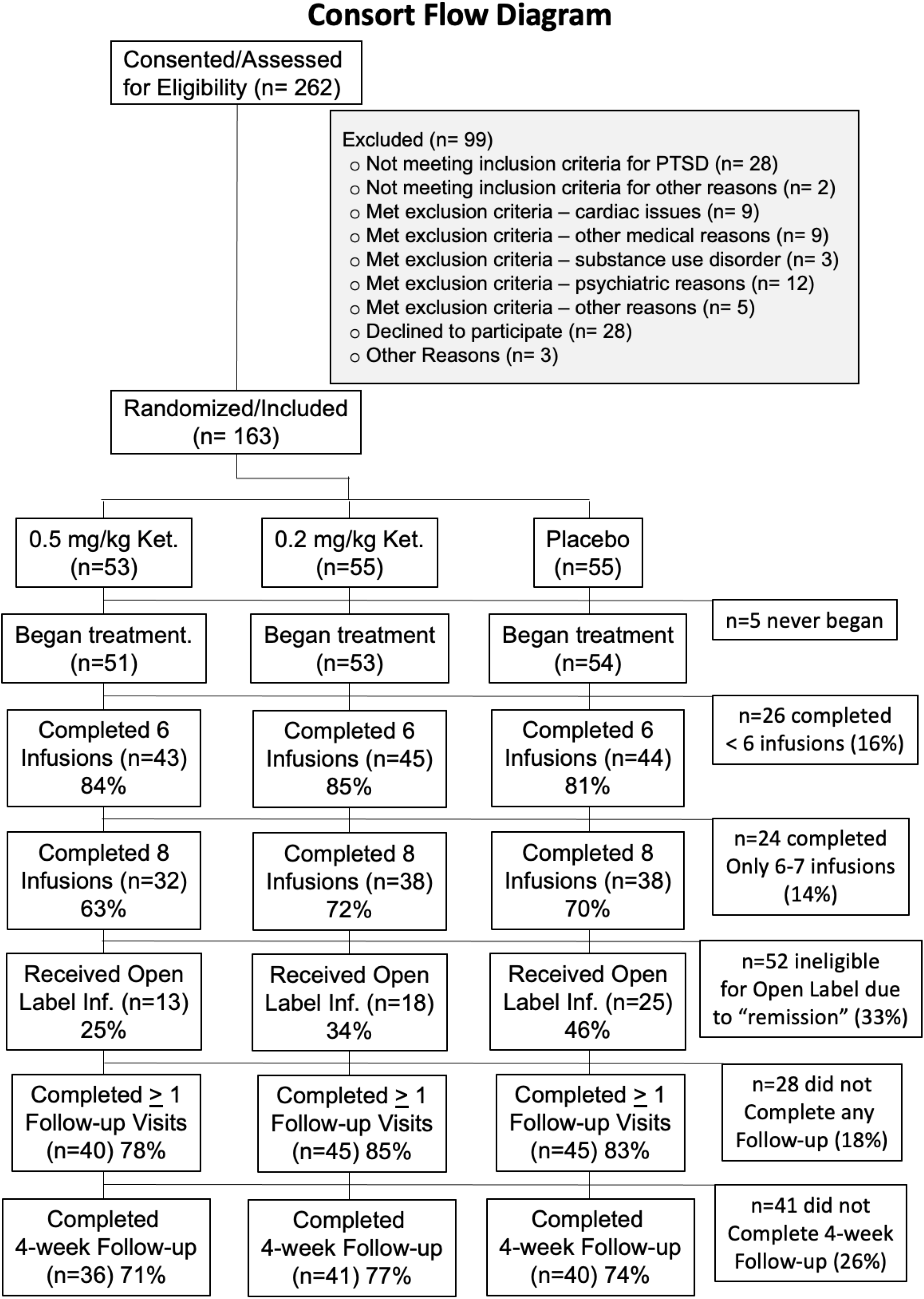

**Table S1.**
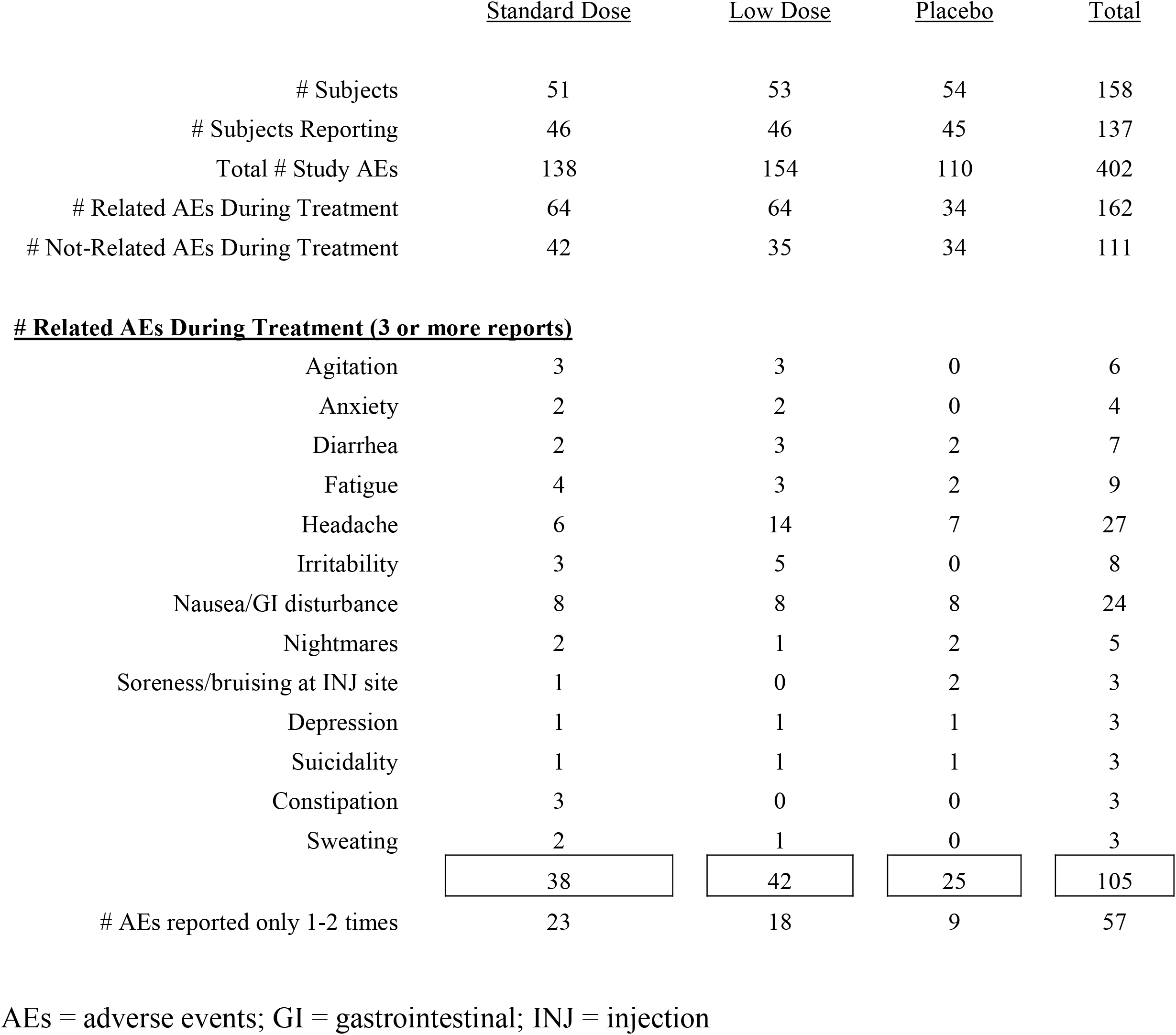
Adverse Events Summary.

|  | <u>Standard Dose</u> | <u>Low Dose</u> | <u>Placebo</u> | <u>Total</u> |
| --- | --- | --- | --- | --- |
| # Subjects | 51 | 53 | 54 | 158 |
| # Subjects Reporting | 46 | 46 | 45 | 137 |
| Total # Study AEs | 138 | 154 | 110 | 402 |
| # Related AEs During Treatment | 64 | 64 | 34 | 162 |
| # Not-Related AEs During Treatment | 42 | 35 | 34 | 111 |
| **# Related AEs During Treatment (3 or more reports)** |  |  |  |  |
| Agitation | 3 | 3 | 0 | 6 |
| Anxiety | 2 | 2 | 0 | 4 |
| Diarrhea | 2 | 3 | 2 | 7 |
| Fatigue | 4 | 3 | 2 | 9 |
| Headache | 6 | 14 | 7 | 27 |
| Irritability | 3 | 5 | 0 | 8 |
| Nausea/GI disturbance | 8 | 8 | 8 | 24 |
| Nightmares | 2 | 1 | 2 | 5 |
| Soreness/bruising at INJ site | 1 | 0 | 2 | 3 |
| Depression | 1 | 1 | 1 | 3 |
| Suicidality | 1 | 1 | 1 | 3 |
| Constipation | 3 | 0 | 0 | 3 |
| Sweating | 2 | 1 | 0 | 3 |
|  | 38 | 42 | 25 | 105 |
| # AEs reported only 1-2 times | 23 | 18 | 9 | 57 |

AEs = adverse events; GI = gastrointestinal; INJ = injection

## REFERENCES

1. Abdallah CG, Averill LA, Akiki TJ, Raza M, Averill CL, Gomaa H, et al. (2019): The Neurobiology and Pharmacotherapy of Posttraumatic Stress Disorder. Annu Rev Pharmacol Toxicol. 59:171–189.

2. Shalev A, Liberzon I, Marmar C (2017): Post-Traumatic Stress Disorder. N Engl J Med. 376:2459–2469.

3. Yehuda R, Hoge CW, McFarlane AC, Vermetten E, Lanius RA, Nievergelt CM, et al. (2015): Post-traumatic stress disorder. Nat Rev Dis Primers. 1:15057.

4. Akiki TJ, Abdallah CG (2018): Are There Effective Psychopharmacologic Treatments for PTSD? J Clin Psychiatry. 80.

5. Krystal JH, Davis LL, Neylan TC, m AR, Schnurr PP, Stein MB, et al. (2017): It Is Time to Address the Crisis in the Pharmacotherapy of Posttraumatic Stress Disorder: A Consensus Statement of the PTSD Psychopharmacology Working Group. Biol Psychiatry. 82:e51–e59.

6. Cipriani A, Williams T, Nikolakopoulou A, Salanti G, Chaimani A, Ipser J, et al. (2017): Comparative efficacy and acceptability of pharmacological treatments for post-traumatic stress disorder in adults: a network meta-analysis. Psychol Med.1–10.

7. Watts BV, Schnurr PP, Mayo L, Young-Xu Y, Weeks WB, Friedman MJ (2013): Meta-analysis of the efficacy of treatments for posttraumatic stress disorder. J Clin Psychiatry. 74:e541–550.

8. Abdallah CG, Sanacora G, Duman RS, Krystal JH (2018): The neurobiology of depression, ketamine and rapid-acting antidepressants: Is it glutamate inhibition or activation? Pharmacol Ther. 190:148–158.

9. Kraus C, Wasserman D, Henter ID, Acevedo-Diaz E, Kadriu B, Zarate CA, Jr. (2019): The influence of ketamine on drug discovery in depression. Drug Discov Today.

10. Feder A, Parides MK, Murrough JW, Perez AM, Morgan JE, Saxena S, et al. (2014): Efficacy of intravenous ketamine for treatment of chronic posttraumatic stress disorder: a randomized clinical trial. JAMA psychiatry. 71:681–688.

11. Feder A, Costi S, Rutter SB, Collins AB, Govindarajulu U, Jha MK, et al. (2021): A Randomized Controlled Trial of Repeated Ketamine Administration for Chronic Posttraumatic Stress Disorder. Am J Psychiatry.appiajp202020050596.

12. Albott CS, Lim KO, Forbes MK, Erbes C, Tye SJ, Grabowski JG, et al. (2018): Efficacy, Safety, and Durability of Repeated Ketamine Infusions for Comorbid Posttraumatic Stress Disorder and Treatment-Resistant Depression. J Clin Psychiatry. 79.

13. Dai D, Lacadie CM, Holmes SE, Cool R, Anticevic A, Averill C, et al. (2020): Ketamine Normalizes the Structural Alterations of Inferior Frontal Gyrus in Depression. Chronic Stress. 4:2470547020980681.

14. Dadabayev AR, Joshi SA, Reda MH, Lake T, Hausman MS, Domino E, et al. (2020): Low Dose Ketamine Infusion for Comorbid Posttraumatic Stress Disorder and Chronic Pain: A Randomized Double-Blind Clinical Trial. Chronic Stress. 4:2470547020981670.

15. Abdallah CG, Roache JD, Averill LA, Young-McCaughan S, Martini B, Gueorguieva R, et al. (2019): Repeated ketamine infusions for antidepressant-resistant PTSD: Methods of a multicenter, randomized, placebo-controlled clinical trial. Contemporary clinical trials. 81:11–18.

16. Singh JB, Fedgchin M, Daly EJ, De Boer P, Cooper K, Lim P, et al. (2016): A Double-Blind, Randomized, Placebo-Controlled, Dose-Frequency Study of Intravenous Ketamine in Patients With Treatment-Resistant Depression. Am J Psychiatry. 173:816–826.

17. Guina J, Rossetter SR, D. RB, Nahhas RW, Welton RS (2015): Benzodiazepines for PTSD: A Systematic Review and Meta-Analysis. J Psychiatr Pract. 21:281–303.

18. Duek O, Spiller TR, Pietrzak RH, Fried EI, Harpaz-Rotem I (2020): Network analysis of PTSD and depressive symptoms in 158,139 treatment-seeking veterans with PTSD. Depress Anxiety.

19. Schiavone FL, McKinnon MC, Lanius RA (2018): Psychotic-Like Symptoms and the Temporal Lobe in Trauma-Related Disorders: Diagnosis, Treatment, and Assessment of Potential Malingering. Chronic Stress (Thousand Oaks). 2:2470547018797046.

20. Krystal JH, Karper LP, Seibyl JP, Freeman GK, Delaney R, Bremner JD, et al. (1994): Subanesthetic effects of the noncompetitive NMDA antagonist, ketamine, in humans. Psychotomimetic, perceptual, cognitive, and neuroendocrine responses. Archives of general psychiatry. 51:199–214.

21. Murrough JW, Iosifescu DV, Chang LC, Al Jurdi RK, Green CE, Perez AM, et al. (2013): Antidepressant efficacy of ketamine in treatment-resistant major depression: a two-site randomized controlled trial. Am J Psychiatry. 170:1134–1142.

22. Fava M, Evins AE, Dorer DJ, Schoenfeld DA (2003): The problem of the placebo response in clinical trials for psychiatric disorders: culprits, possible remedies, and a novel study design approach. Psychother Psychosom. 72:115–127.

23. Mohamed S, Johnson GR, Sevilimedu V, Rao SD, Hicks PB, Chen P, et al. (2020): Impact of Concurrent Posttraumatic Stress Disorder on Outcomes of Antipsychotic Augmentation for Major Depressive Disorder With a Prior Failed Treatment: VAST-D Randomized Clinical Trial. J Clin Psychiatry. 81.

24. Fava M, Freeman MP, Flynn M, Judge H, Hoeppner BB, Cusin C, et al. (2020): Double-blind, placebo-controlled, dose-ranging trial of intravenous ketamine as adjunctive therapy in treatment-resistant depression (TRD). Mol Psychiatry. 25:1592–1603.

25. Su TP, Chen MH, Li CT, Lin WC, Hong CJ, Gueorguieva R, et al. (2017): Dose-Related Effects of Adjunctive Ketamine in Taiwanese Patients with Treatment-Resistant Depression. Neuropsychopharmacology. 42:2482–2492.

